# Where to learn to flatten the curve: a modelling study

**DOI:** 10.1101/2021.03.23.21254166

**Authors:** Matthew J Silk, Simon Carrignon, R. Alexander Bentley, Nina H Fefferman

## Abstract

**Background:** Individual behavioural decisions are responses to both a person’s perceived social norms and could be driven by both their physical and social environment. In the context of the COVID-19 pandemic, these environments correspond to epidemiological risk from contacts and the social construction of risk by communication within networks of friends. Understanding when, and under which circumstances, each modality of influence can foster the widespread adoption of protective behaviours is critical for shaping useful, practical public health messaging that will best enhance the public response.

**Methods:** We use a multiplex network approach to explore how information from both physical contact and social communication networks is driving a mitigating behavioural response to disease risk.

**Findings:** We show that maintaining focus on awareness of risk in each individual’s physical layer contacts promotes the greatest reduction in disease spread, but only when an individual is aware of the symptoms of a non-trivial proportion of their physical contacts (approximately 20% or more). Information from the communication layer was less useful when these connections matched less well with physical contacts and contributed little in combination with accurate information from the physical layer.

**Interpretation:** We conclude that maintaining social focus on local outbreak status will allow individuals to structure their perceived social norms appropriately and respond more rapidly when risk increases. Finding ways to relay accurate local information from trusted community leaders could improve mitigation even where more intrusive/costly strategies, such as contact-tracing, are not possible.

## Introduction

Our current best public health recommendations for mitigation of the COVID-19 pandemic rely on using behavioural interventions such as social distancing and mask wearing, and behaviourally driven acceptance of vaccines (where available) to curtail transmission of infection. The success of these policies requires widespread adherence to achieve epidemic control; as with herd immunity, threshold effects in efficacy mean that gaps in adoption can quickly compromise any benefits^[1,2]^. Therefore, identifying how the adoption of these behaviours is shaped over the course of an epidemic is a key challenge in designing effective mitigation strategies^[3-6]^.

Adherence, however, relies on individual behavioural choices and so can be complicated to understand and predict^[3,7]^f. Well-established theory from psychology acknowledges that the factors influencing whether or not people take action are complicated^[8,9]^. One of the dominant theories (the theory of planned behaviour^[10]^), posits that action is the composite result of the individual’s attitudes and beliefs, the individual’s perception of social norms regarding that behaviour, and the individual’s perception of their own behavioural control over their actions (alternative theories of behaviour, such as Value-Belief-Norm theory^[11]^ also posit similar influences, though in different relation to each other). In the case of COVID-19, adoption of and adherence to behavioural interventions are therefore likely to be predicated on perception of two main features: a) individual attitudes and beliefs about personal risk of infection and its consequences^[12]^, and b) the social norms around adherence in the individual’s community^[13]^.

An individual’s perception of these features is shaped by communication within their network of friends, neighbours, and community leaders^[14,15]^. Most likely, the network of a person’s close physical contacts, through which they risk infection, differs from their regular communication network (in person and online) of people who contribute to their attitudes and beliefs surrounding preventative behaviours, and from whom they are likely to estimate the social norms of their community. These distinct networks underlie a disconnect between someone’s perception of their risk versus their actual risk. On one hand, an individual’s communication network could provide early warning of encroaching exposure risks derived from the spread of awareness ahead of the infection itself ^[16,17]^. On the other hand, the mismatch between the communication layer and infection layer may mean that an individual could underestimate their risk (e.g., communication networks are likely to be more sparsely connected than networks of infection-relevant contacts in populations that are not socially distancing). Despite this, we still understand relatively little about the potential implications of acquiring information from these two different sets of contacts.

The dynamics triggered by the spread of awareness through the population are further complicated by the timescales of observable risk due to the etiology of COVID-19. The latency in the development of symptoms and the capacity for presymptomatic, or even asymptomatic, transmission make estimation of real-time risk by surveillance complicated, even without considering different sources of information^[18]^. In terms of understanding disease prevalence, the relative reliance of individuals in shaping their beliefs, and thus their actions, on their own direct observation of health among their daily physical contact network may have an effect that is distinct from that of their (potentially more geographically distant) communication network. The balance of these distinct network effects may therefore be the critical feature in determining the success of behavioural public health measures to combat COVID-19.

We employ a multiplex network method to test the relative adoption of behavioural interventions in populations of individuals who rely on a) their communication network layer only (henceforth referred to as simply the “communication layer”), b) their physical contact network layer only (henceforth referred to as the “infection layer”), and c) both layers simultaneously to inform their understanding of COVID-19, and therefore their individual adherence to protective behaviours such as mask wearing or social distancing. We further consider the influence of structure in both layers of the network and how that structure might impact the behaviour of populations as they rely on perceptions constructed from contacts in those layers. Geographic and social heterogeneity in contact structure are modelled at low and high levels of modularity (i.e. differences in local versus global density of contacts; see Methods). In addition to homogeneous mixing, we consider homophily based upon predisposition, which are modelled as social norms that can be shared in the communication network. While not exhaustive, these studies offer insight into how communities can reinforce the types of informational access that foster protective behavioural decision making among their members.

## Methods

### Overview

We used stochastic models to test how the awareness of symptomatic neighbours in either a) the set of people that a person who communicates with on a regular basis (their communication layer), b) the set of people that a person is in close proximity to (their infection layer), or c) both of these sets of contacts can impact epidemic spread of an infection with COVID-19 like dynamics. We simulated realistic, multiplex social networks for our populations that coupled a layer of infection-relevant contacts through which the epidemic was simulated and a communication layer through which concern about the disease could simultaneously spread. All modelling was conducted in R3.6.1^[19]^ and the code used is provided on GitHub (https://github.com/matthewsilk/CoupledDynamics2_layeruse). The general modelling framework was the same as that used by Silk et al.^[20]^ and is described in that paper and in the Supplementary Material.

### Population generation

We generated populations of 2000 individuals (a balance between minimising stochasticity in early epidemic outcomes and computational efficiency), which consisted of children (24%), young adults (63%) and old adults (13%). Age classes could differ in the social connections, epidemiological outcomes and concern about the disease (as detailed below). Individuals also had one of two baseline predispositions and homophily by predisposition impacted patterns of social connections (in either or both layers of the multiplex network).

### Social network generation

We used the same 9 multiplex social networks as detailed in Silk et al.^[20]^. These were coupled, multiplex networks that connected all individuals within a communication layer that influenced the spread of concern about the disease and an infection layer that influenced the transmission of the pathogen itself. A full description of the algorithm used to generate these networks is provided in the Supplementary Material. For this study, global edge densities were always higher in the infection layer than in the communication layer. The network contained either a) no homophily in either layer, b) homophily in the communication layer, or c) homophily in both layers. Community structure was introduced using a re-wiring algorithm (as detailed in the Supplementary Material): either the relative modularity of both layers was set to 0·4, both to 0·6, or the infection layer was set to 0·6 and the communication layer to 0·4. Each child was assigned two parents from the same predisposition and community. If children shared one parent they also shared the other but parents could be connected or unconnected. Each young adult formed connections with a number of old adults of the same predisposition (representing older relatives, friends or community members) as detailed in the Supplementary Material. Children shared the same connections to old adults as their parents. When the multiplex network was constructed we re-assigned parents from the infection layer to match those in the communication layer. Child-old adult connections were re-assigned accordingly.

### Concern model

We used the same concern model as Silk et al.^[20]^. Concern about the disease was modelled as a complex contagion^[21]^ through the communication layer. Whether an individual was adherent to mitigation measures or not (a binary trait) was based on a Bernoulli draw in which the probability of adherence depended on an underlying trait continuous we term concern. As a result, individuals could fluctuate between adherent or non-adherent states and this was more likely if they had intermediate values of concern. Concern could be influenced by a) Social Construction (becoming more concerned if neighbours in the communication layer were adherent), b) Reassurance (becoming less concerned if all neighbours in the communication layer were perceived to be healthy (i.e. symptom free) and c) Awareness (becoming more concerned if network neighbours were symptomatic). For this study the information gained for Awareness could be gained from either the communication layer, the infection layer or both layers. Because an individual is unlikely to find out about the status of every individual in their infection layer, we conducted additional simulations in which there was imperfect detection of symptomatic contacts in the infection layer (probability of detection: 0.5, 0.2 and 0.05). The concern of children was not modelled. They were assigned as adherent if either or both of their parents were concerned.

Each time an individual became adherent they cut connections to a negligible edge weight (see Supplementary Material) within the infection layer while maintaining their connectivity in the communication layer. If an individual became non-adherent then these edge weights returned to their full weight.

### Infectious disease model

Our infectious disease model is the same stochastic network model described in Silk et al.^[20]^ with parameter values adapted from^[22, 23]^. The model contains susceptible (S), exposed (E), pre-symptomatic (I1), symptomatic (I2), hospitalised (I3), recovered (R) and dead (D) compartments. Parameter values are provided in Table S1 and details of the algorithm used are provided in the Supplementary Material.

### Simulations

For this paper we conducted simulations for the nine multiplex networks described (with different combinations of homophily by predisposition and modularity), for 50 values of the Social Construction and Reassurance effects (paired draws from independent uniform distributions) and 10 values of the Awareness effect. We then conducted simulations in which the Awareness effect (observational learning of symptomatic infection) applied to a) contacts in the communication layers, b) contacts in the infection layer and c) contacts from both layers combined. For scenario b) we repeated the full set of simulations with 0·5, 0·2 and 0·05 probability of symptomatic contacts being detected at each day. This resulted in a total of 27,000 independent simulation runs. For each simulation run we simulated a time period of 300 days. The simulation algorithm was the same used in Silk et al.^[20]^ and is detailed in the Supplementary Material.

### Analysis

To compare between different runs of the simulations we quantified the height of the epidemic peak at a population level by aggregating the daily counts of symptomatic infections in all 10 communities. This measure of the height of the epidemic peak indicated how successfully each simulated population managed to “flatten the curve” with their adherence to mitigating behaviours^[4]^. We compare epidemic peaks from when individuals learned about symptomatic network neighbours from different types of social contact while considering values of the Social Construction and Reassurance effects. To help explain some of the differences between the infection and communication layers in their ability to “flatten the curve” we also examined the similarity of connections in these layers by quantifying the proportion of contacts in each layer that were also present in the other for each multiplex network.

## Results

When we assume individual can identify 100% of symptomatic contacts the Awareness Effect is more effective in flattening the curve when people respond to illness in their infection layer rather than in their communication layer (panels a versus b in Figs. 1 and 2). When this is the case, even relatively weak Awareness Effects can contribute to flattening the curve. Using information from the infection layer is nearly as effective as using information from both the infection and communication layers except when the Awareness Effect is very weak (compare panels b and c in Figs 1 and 2). When social construction is weak there is an important difference regardless of the strength of the Reassurance effect (Fig. 1), while when social construction is strong it plays an important role in flattening the curve except in the case when the Reassurance Effect is strong, meaning that differences caused by the source of information for the Awareness Effect are only noticeable when this is the case (Fig. 2).

**Figure 1.**
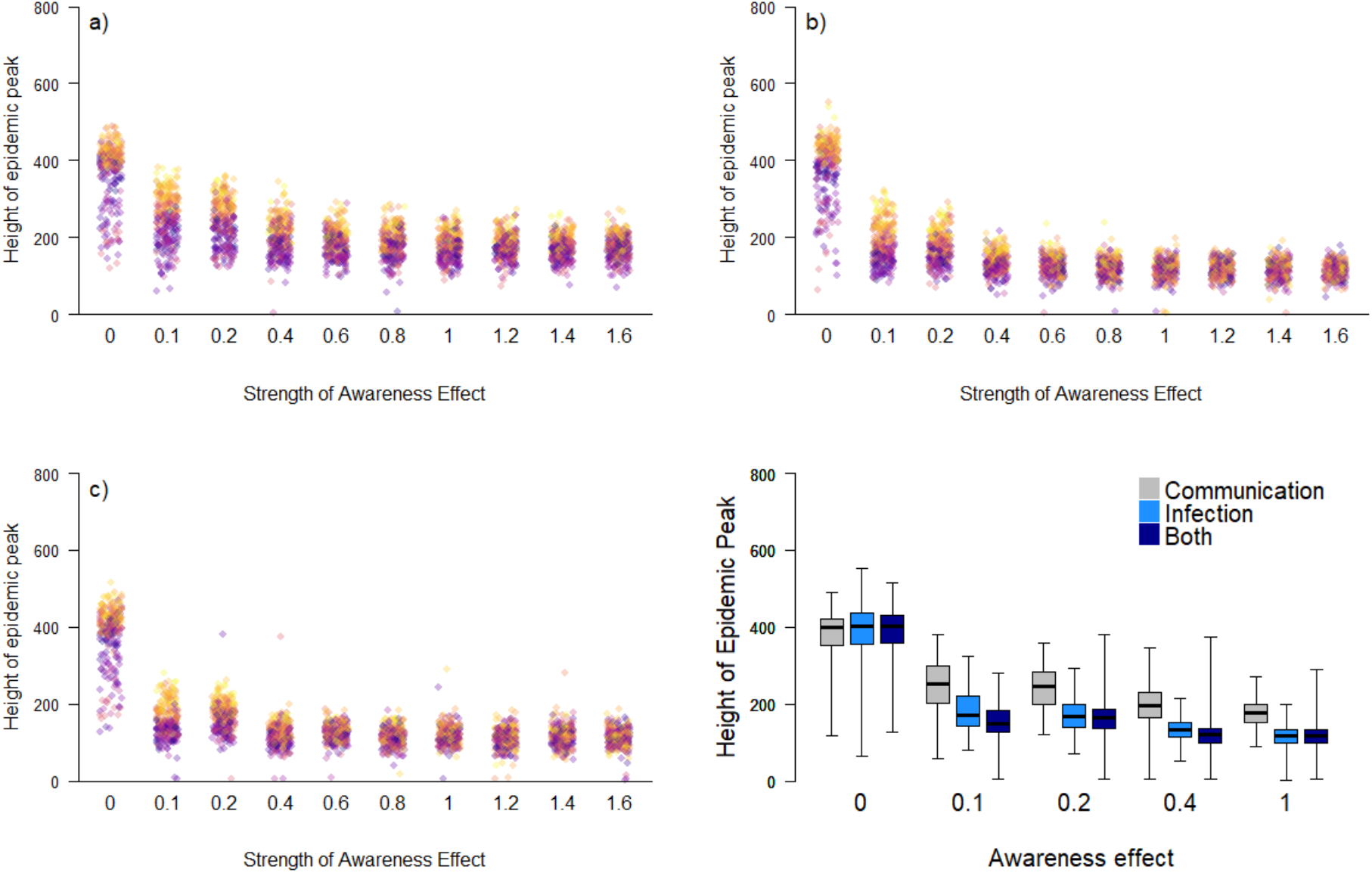
The relationship between the height of the epidemic peak and strength of the Awareness effect when Social Construction is weak (<0.3). An individual learns of symptomatic contacts from a) their communication layer, b) their infection layer (learning from 100% of contacts) and c) both layers together. The colour of points in panels (a-c) indicates the strength of the Reassurance effect: yellow indicating a strong reassurance effect through to purple indicating a weak Reassurance effect. In panel (d) we contrast the height of the epidemic peak directly for selected values of the Awareness effect. Boxes indicate the interquartile range, the bold horizontal line the median and the whiskers extend to the full range of the data.

**Figure 2.**
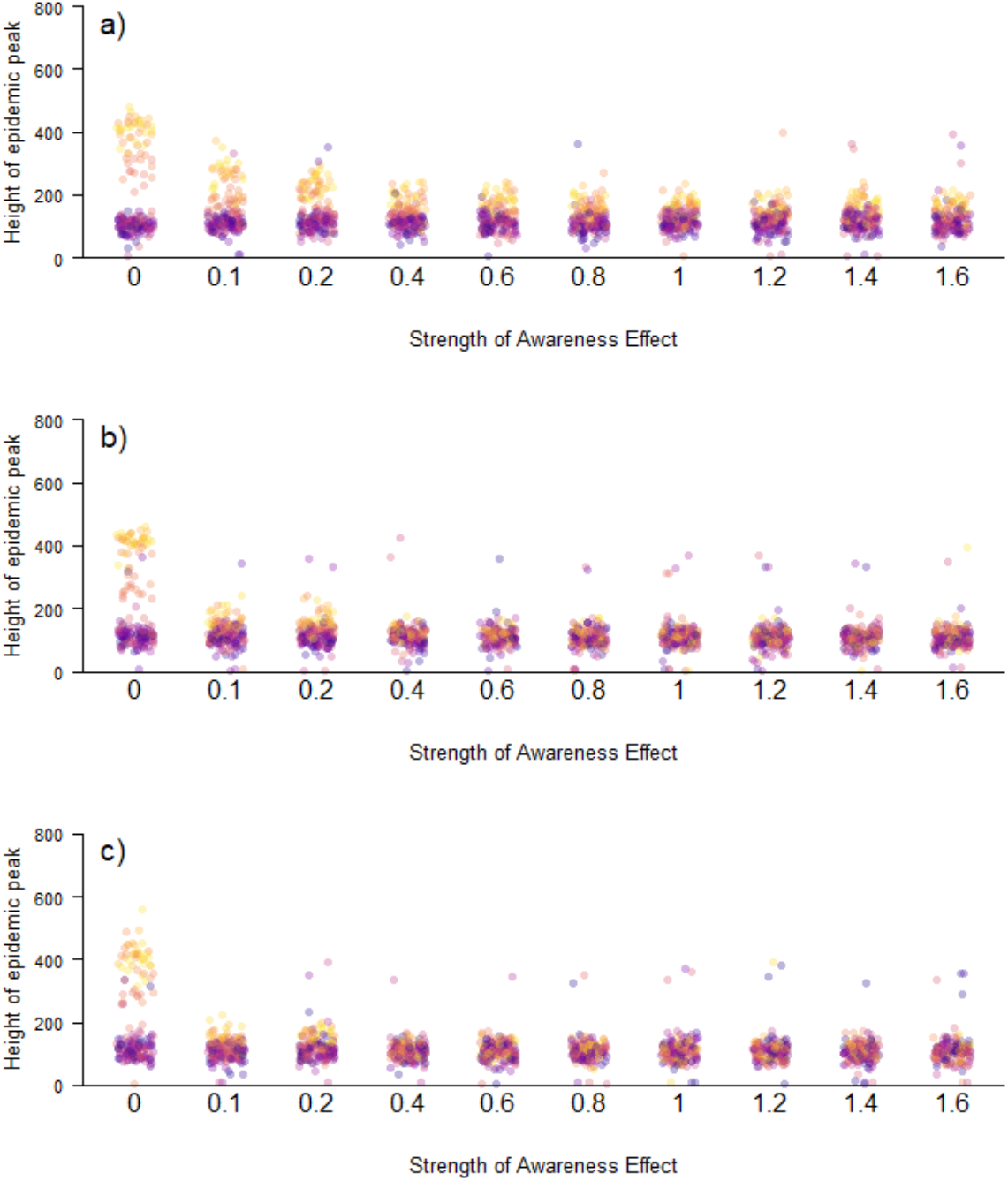
The relationship between the height of the epidemic peak and strength of the Awareness effect when Social Construction is strong (>0.3). An individual learns of symptomatic contacts from a) their communication layer, b) their infection layer (learning from 100% of contacts) and c) both layers together. The colour of points indicates the strength of the Reassurance effect: yellow indicating a strong reassurance effect through to purple indicating a weak Reassurance effect.

Consequently, we focus on the case when Social Construction is weak for subsequent results. A second notable difference that arises when people respond to prevalence in their infection layer rather than the communication layer is that the strength of the Reassurance Effect becomes less important. When individuals respond to illness in their communication layer, the epidemics always have higher peaks with a strong Reassurance effect (also reliant on the communication layer) even when the Awareness effect is strong and the curve has been flattened (Fig. 1a). However, when the Awareness Effect is stronger (>0·6), learning about illness from the infection layer or both layers results in similar epidemic peaks regardless of the strength of the Reassurance Effect (Figs. 1b and 1c).

When we assume that individual can partially identify the symptomatic contacts in their infection layer, the mitigating effect is reduced considerably in our networks (Fig. 3). When there is a 50% chance of an individual detecting an ill neighbour in their infection layer, the height of the epidemic peak remains lower than when an individual gains information on the prevalence of infection from their communication layer with the difference increasing as the strength of the Awareness Effect gets stronger. When there is a 20% chance of detection in the infection layer, the epidemic peak is marginally higher than when (accurate) information is used from the communication layer with a weak Awareness Effect and slightly lower with a strong Awareness Effect. When there is a 5% chance of detection the mitigating influence of the Awareness Effect is very limited indeed and restricted to strong Awareness Effects. The structure of the network was relatively unimportant in determining the success with which populations were able to “flatten the curve” (Fig. 4, Fig. S1). Most strikingly, there was a small negative impact on the value of information from the communication layer when there was homophily only in that layer and not in the infection layer (networks 7-9). When this was the case epidemic peaks remained higher when individuals acquired information on local prevalence from their communication layer. This pattern was driven by their being a greater mismatch between the two layers when there was only homophily in the communication layer; a lower proportion of edges in the infection layer were also present in the communication layer (Fig. 5). It is harder, therefore, to flatten the curve when key aspects of structure of communication and infection layers are mismatched and individuals gain information on illness from their communication layer. Otherwise there were no clear and consistent patterns related to network structure over the range of structures tested here. Results were qualitatively similar regardless of the strength of the Awareness Effect (Fig. S1).

**Figure 3.**
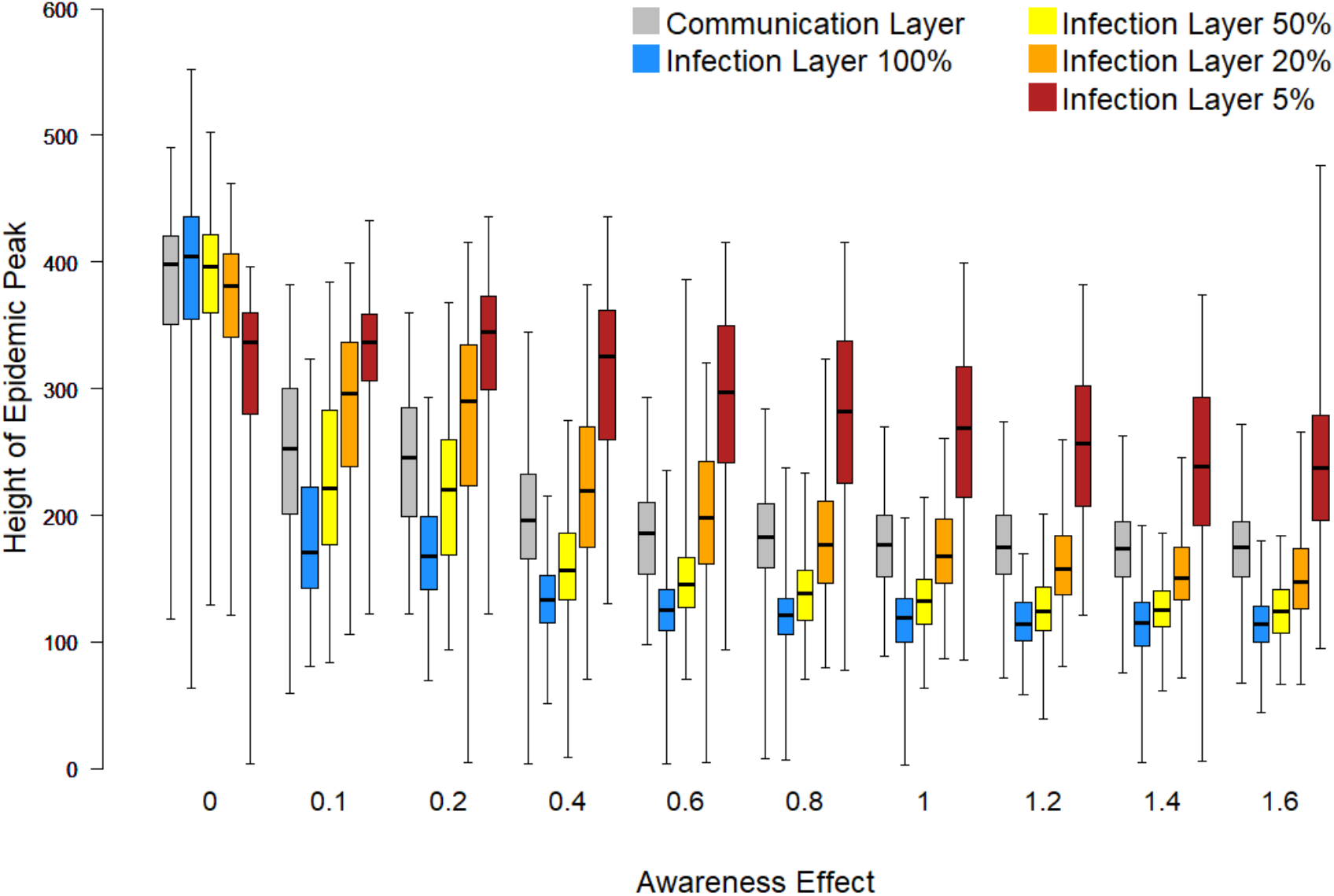
The relationship between the height of the epidemic peak and strength of the Awareness effect when Social Construction is weak (<0.3). We show the relationship when Awareness is acquired through the communication layer (grey) and the infection layer when 100% (blue), 50% (yellow), 20% (orange) and 5% (red) of symptomatic contacts are detected each day. Boxes indicate the interquartile range, the bold horizontal line the median and the whiskers extend to the full range of the data.

**Figure 4.**
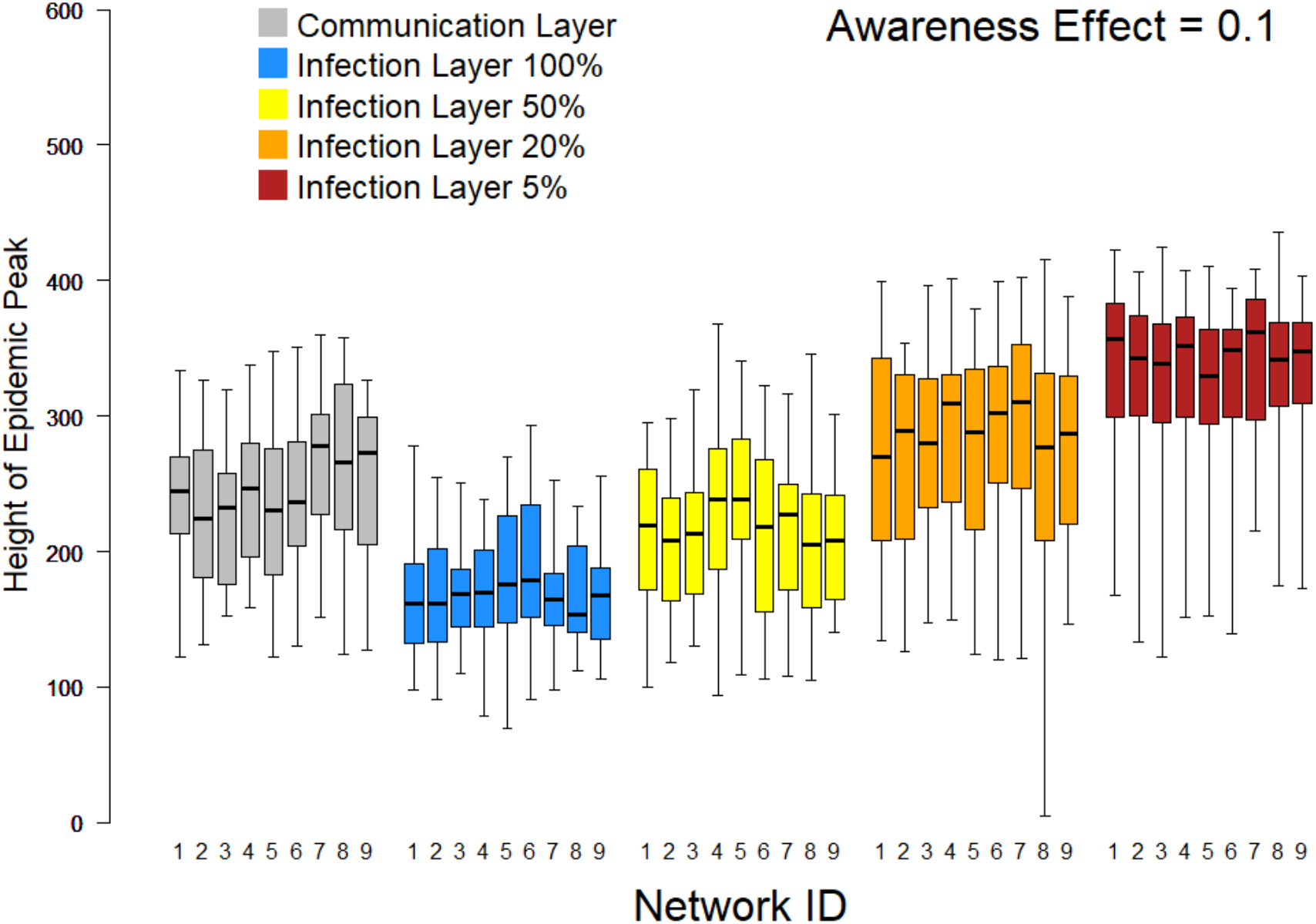
The relationship between the height of the epidemic peak and how an individual finds out about symptomatic contacts when the Social construction effect is weak (<0.3) for an Awareness effect of 0.1 plotted separately for each of the nine multiplex networks used in the study. Networks 1-3 have no homophily in either layer, networks 4-6 have homophily in both layers and networks 7-9 have homophily in the communication layer only. In networks 1, 4 and 7 both layers have a relative modularity of 0·4, in networks 2, 5 and 8 both layers have a relative modularity of 0·6, and in networks 3, 6 and 9 the relative modularity of the infection layer is 0·6 and the relative modularity of the communication layer is 0·4. Plots for other Awareness effects our qualitatively similar (see Fig. S1).

**Figure 5.**
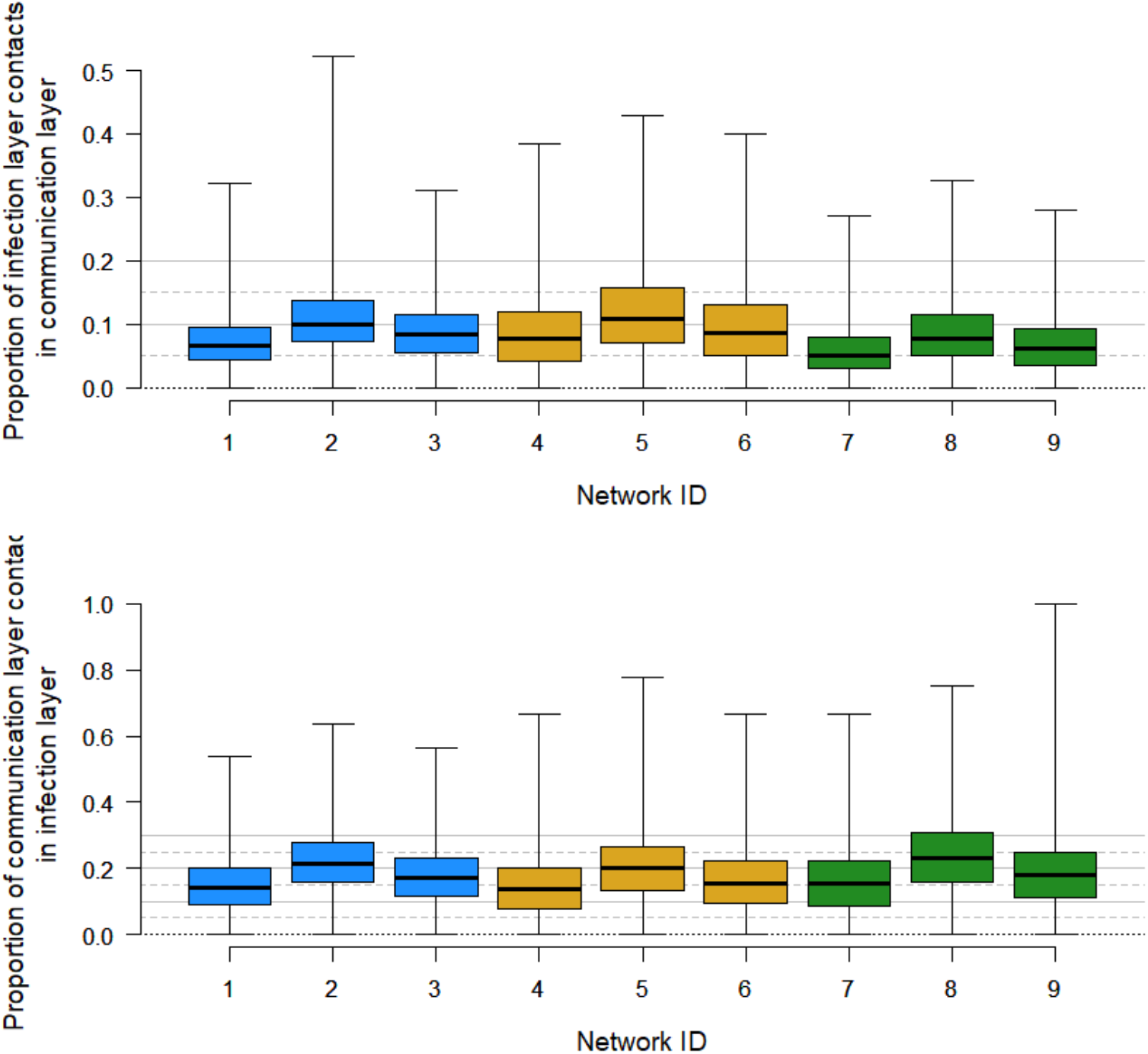
The proportion of contacts in each layer of the multiplex network that are present in the other layer. Panel (a) shows the proportion of infection layer contacts also present in the communication layer and panel (b) show the proportion of communication layer contacts also present in the infection layer. Networks 1-3 have no homophily in either layer, networks 4-6 have homophily in both layers and networks 7-9 have homophily in the communication layer only. In networks 1, 4 and 7 both layers have a relative modularity of 0·4, in networks 2, 5 and 8 both layers have a relative modularity of 0·6, and in networks 3, 6 and 9 the relative modularity of the infection layer is 0·6 and the relative modularity of the communication layer is 0·4.

## Discussion

Despite the natural inclination to believe that the influence of community leaders and trusted social contacts can help us each to make better decisions, in this case, the results of our study demonstrate that the impact of that trusted community may instead compromise our ability to respond to the actual risks around us (see Figs. 1,2, and 3) if the alternative is accurate information on prevalence in an individual’s likely physical contacts. However, the advantage brought about by an individual’s knowledge of the prevalence in their “infection layer” declines very rapidly as the accuracy of this information deteriorates. This has strong, direct implications for individuals living in circumstances in which their physical contacts are likely removed from their social spheres of influence. Critically, this pattern reflects large urban centres in which individuals may physically contact may people using public transportation, or riding elevators and moving among offices or apartments in large in high-rise buildings, but are likely instead to rely on a mostly separate community of family, faith, or recreational activities for social community and conversation from which they will form their perceptions of risks and norms.

Of course, this result relies on the low overlap between contacts present in both the communication and disease layers that shape an individual’s perceptions and risks (see Fig. 5). If those layers were instead identical (as in small, remote rural communities or highly segregated, small, well-mixed communities as exist affiliated with some religious groups), then the communication layer and infection layer will be equivalent in the information they provide, meaning that more information is available on local prevalence in an individual’s infection layer and so improving the decision-making of individuals based on observations of their personal networks.

Our results show that the availability of accurate information from an individual’s infection layer is much more effective in mitigating disease spread (i.e. “flattening the curve”) than using only their communication layer, and that when this is the case using information from both only performs marginally better than using the infection layer alone. However, as the availability of accurate information from the infection layer declines the success of mitigation declines very rapidly. In our networks, a >20% chance of detecting each symptomatic neighbour in the infection layer is required for mitigation to be more successful than when individuals use their communication layer alone.

The former result suggests that populations comprised of individuals who tend more towards independent risk assessment than on reliance on community leadership may respond better to public health interventions. However, the latter result indicates the importance of highly accurate information from the infection layer at a community level. Therefore any social norms that reduce observability of infection in a local community can undercut the efficacy of recommended behavioural interventions. This is especially important in shaping public messaging since both within group density (i.e. modularity) and closeness of beliefs within a community (i.e. homophily) have less of an impact than the information on which the members of that community rely (see Fig. 4). We therefore strongly support the adoption of public reports of identified cases in local communities that come into regular potential contact with each other. While, of course, this requires sensitivity to personal privacy, regular announcements/reminders at a city, company, school, or neighbourhood level of active disease prevalence can potentially provide critical and effective reinforcement for the individual adoption of behaviours that can protect everyone.

Awareness itself is not without complexity. The centralized collection and analysis of data at regional or national scales involves logistical challenges and can cause delay in reporting that information back to the public^[24]^. It is also frequently the case that communities pay more attention to, and place greater trust in, local sources of information than in more remote sources^[25]^. Policies that focus on community leadership to ensure a local focus for awareness helps to address both of these difficulties.

Perhaps most importantly, this study suggests the need for leaders of social groups to ensure attention is paid to cases of COVID-19 in their community. Luckily, this is in keeping with the mission of many social groups focused on community support. Communities of worship, social action organizations, and community volunteer groups have all been active participants across the globe in making sure that individuals who are unwell but not so severely impacted as to be hospitalized have access to groceries, medicines, and wellness checks. By actively highlighting the need for these services within their own community, these actions themselves support broader adoption of preventative behaviours and thereby not only help individuals already affected by COVID-19, but actively decrease the likely magnitude of local impacts from the pandemic.

## Conclusion

One of the most fundamental challenges in creating effective public health policies is the design of recommendations that will not only achieve theoretical outcomes but will be adopted by enough of a willing public to accomplish those outcomes in the real world. Integrating an understanding of how individual perceptions shape behaviours, and how social context itself shapes perceptions has become one of the critical stumbling points in our local, national, and global response to the COVID-19 pandemic. Our results clearly show that local, accurate, rapid, and trusted information can enable better emergent behaviours. Thankfully, these paths are within the capability of our public health community and local community leadership to provide.

### Data Sharing Statement

No primary data was collected as part of this effort. All relevant code is publicly available at (https://github.com/matthewsilk/CoupledDynamics2_layeruse).

## Supplementary Methods

### Overview

We used stochastic models to test how the awareness of symptomatic neighbours in either a) the set of people that a person who communicates with on a regular basis (their communication layer), b) the set of people that a person is in close proximity to (their infection layer), or c) both of these sets of contacts can impact epidemic spread of an infection with COVID-19 like dynamics. We simulated realistic, multiplex social networks for our populations that coupled a layer of infection-relevant contacts through which the epidemic was simulated and a communication layer through which concern about the disease could simultaneously spread. All modelling was conducted in R3.6.1^[19]^ and the code used is provided on GitHub (https://github.com/matthewsilk/CoupledDynamics2_layeruse). The general modelling framework was the same as that used by Silk et al.^[20]^.

### Population generation

We generated populations of 2000 individuals (a balance between minimising stochasticity in early epidemic outcomes and computational efficiency), which consisted of children (24%), young adults (63%) and old adults (13%). These proportions were set to match recent US demographic data. Age classes could differ in the social connections, epidemiological outcomes and concern about the disease (as detailed below). Individuals also had one of two baseline predispositions, with 50% of each predisposition. Homophily by predisposition impacted only patterns of social connections (in either or both layers of the multiplex network).

### Social network generation

We generated 9 multiplex social networks as detailed in Silk et al.^[20]^. These were coupled, multiplex networks that connected all individuals within a communication layer that influenced the spread of concern about the disease and an infection layer that influenced the transmission of the pathogen itself. Briefly, networks were generated as follows:

1. For each layer, within age class and within predisposition connections were simulated as Erdös-Renyi random graphs using igraph^[26]^. We specified age-specific edge densities for within predisposition connections. For this study, global edge densities were always higher in the infection layer than in the communication layer.
2. Between predisposition connections were then added to form three overall networks for each age class in each layer. We specified age-specific between-predisposition edge densities (which could differ from within-predisposition densities). The level of homophily according to predisposition is defined by the difference between within- and between-predisposition edge densities. We included three types of homophily in the multiplex network: a) no homophily in either layer, b) homophily in the communication layer, or c) homophily in both layers.
3. We then introduced community structure into each network layer. A rewiring algorithm was used to impose community structure within the three age-class networks as per Silk et al.^[20]^. We implemented a block model with 10 communities and re-assigned edges to achieve a target modularity while maintaining the initial edge density and level of homophily. We used 0·4 and 0·6 as target relative modularity^[27]^ for our community structure. The proportion of each predisposition within each community was the same as the overall population. In this study either both layers had a modularity of 0·4, both had a modularity of 0·6, or the infection layer had a modularity of 0·6 and the communication layer a modularity of 0·4 (as per Silk et al.^[20]^).
4. Each child was assigned two parents from the same predisposition and community. If children shared one parent they also shared the other but parents could be connected or unconnected.
5. Each young adult formed connections with a number of old adults of the same predisposition (representing older relatives, friends or community members). The number of connections for each young adult was a drawn from a Poisson distribution (..=3). Connections could occur within or between communities with the probability of within-community connections being the same as the modularity of the network. Children shared the same connections to old adults as their parents.

We used 8 different simulated networks in total for this study (four for the communication layer and four for the infection layer), which when combined generated nine different multiplex networks that were a full combination of the three homophily conditions and three modularity conditions defined above. When the multiplex network was constructed we re-assigned parents from the infection layer to match those in the communication layer. Child-old adult connections were re-assigned accordingly.

### Concern model

We used the same concern model as Silk et al.^[20]^. Concern about the disease was modelled as a complex contagion^[21]^ through the communication layer. Whether an individual was adherent to mitigation measures or not (a binary trait) was based on a Bernoulli draw in which the probability of adherence depended on an underlying trait continuous we term concern. As a result, individuals could fluctuate between adherent or non-adherent states and this was more likely if they had intermediate values of concern. Concern could be influenced by the following factors with changes modelled on a logit scale.

1. For this study all adults started with the same concern. The initial level of concern was defined with an expectation of 20% of adults being adherent to mitigation measures at the start of each simulation run.
2. Social Construction of concern – concern could change based on the proportion of immediate neighbours in the communication layer of the network that were adherent in the previous day. We modelled this change as a linear increase in belief as the proportion of adherent connections increased. We allowed the strength of this relationship to vary, drawing 50 different values from a uniform distribution between 0 and 0·5 for the effect size per day. This range of parameter values were selected based on prior use of the model (Silk et al.^[20]^).
4. Reassurance effect from communication neighbours – concern could be reduced at each day that all of an individual’s connections in the communication layer were symptom free (whether susceptible, exposed, pre-symptomatic or recovered). We drew 50 values from a uniform distribution between −0·1 and 0 (which we paired with equivalent values of the Social Construction effect).
5. Awareness effect based on prevalence of symptomatic contacts – concern could increase based on the number of an individual’s connections that were symptomatic. We assigned 10 values (0, 0·1, 0·2, 0·4, 0·6, 0·8, 1, 1·2, 1·4 and 1·6) for this per day increase in concern per symptomatic neighbour. For this paper the awareness effect could depend on a) an individual’s contacts in the communication layer, b) an individual’s contacts in the infection layer or c) an individual’s contacts in both layers combined. Because an individual is unlikely to find out about the status of every individual in their infection layer, we conducted additional sets of simulations in which there was imperfect detection of symptomatic contacts in the infection layer. We used detection probabilities of 50%, 20% and 5%.
6. The concern of children was not modelled. They were assigned as adherent if either or both of their parents were adherent. Each time an individual became adherent they cut connections within the infection layer while maintaining their connectivity in the communication layer. Individuals cut connections with a probability of 0·5, with edges cut to have edge weights of 0·001 (i.e. not removed but sufficiently low to result in a negligible probability of transmission). If an individual became non-adherent then these edge weights returned to their full weight.

### Infectious disease model

Our infectious disease model is the same stochastic network model described in Silk et al.^[20]^ with parameter values adapted from Weitz et al. ^[22]^ and Lofgren et al.^[23]^. The model contains susceptible (S), exposed (E), pre-symptomatic (I1), symptomatic (I2), hospitalised (I3), recovered (R) and dead (D) compartments. The transition between compartments is detailed below and parameter values are provided in Table S1 (reproduced from Silk et al.^[20]^). The infection model proceeded as follows (reproduced from Silk et al.^[20]^):

1. The transition from susceptible to exposed depended on the number of contacts each susceptible individual had with infected individuals (I1, I2, I3). Each contact was associated with a pre-defined probability of transmission. We selected a value for the transmission probability that resulted in unmitigated epidemics infecting approximately 80% of the population as per Ferguson et al._[28]_.
2. The length, in days, of the incubation period (exposed; E) and of each infectious period (I1-3) were set using draws from Poisson distribution (Table S1).
3. Individuals could transition from symptomatic (and at home) to hospitalised, and from hospitalised to dead with pre-defined daily probabilities which differed for young (low risk) adults and old (high risk) adults. These daily probabilities were calculated based on Weitz et al. ^[22]^ and Lofgren et al.^[23]^ and then adapted to produce a realistic proportion of cases being hospitalised during trial runs of the simulations (c.f. Reno et al.^[29]^). Children could be symptomatic (I2) but were never hospitalised reflecting the very low probability of this occurring in reality^[30]^. All individuals that reached the end of each infectious period without further progression of the disease were deemed to have recovered.
4. Symptomatic (I2) and hospitalised (I3) individuals cut all connections in the infection layer of the network to 0.001. These connections were restored to their full value if and when individuals recovered.

### Simulations

For this paper we conducted simulations for the nine multiplex networks described (with different combinations of homophily by predisposition and modularity), for 50 values of the Social Construction and Reassurance effects (paired draws from independent uniform distributions) and 10 values of the Awareness effect. We then conducted simulations in which the Awareness effect (observational learning of symptomatic infection) applied to a) contacts in the communication layers, b) contacts in the infection layer and c) contacts from both layers combined. For scenario b) we repeated the full set of simulations with 0·5, 0·2 and 0·05 probability of symptomatic contacts being detected at each day. This resulted in a total of 27,000 independent simulation runs. For each simulation run we simulated a time period of 300 days.

The simulations proceeded as follows (reproduced from Silk et al.^[20]^):

1. Ten random individuals were seeded with infection (in the exposed [E] class of the compartmental model).
2. Individuals were allocated initial values of belief based on pre-defined starting values and these were used to calculate which individuals were adherent on the first day.
3. The algorithm repeated a sequence events on each day, namely:
  a. The infection layer of the network was rewired so that adherent individuals cut a 50% of their connections to have negligible edge weight.
  b. The infection layer of the network was rewired so that newly symptomatic individuals cut their connections to have negligible edge weight.
  c. The infection layer of the network was rewired so that newly recovered individuals restored their full connections (edge weights increased to 1)
  d. We recorded the identity of all individuals that were currently symptomatic or hospitalised.
  e. We ran the concern model to update the concern and adherence of individuals.
  f. We ran the infectious disease model to update the infection status of all individuals
4. The algorithm moves on to the next day until the end of the defined time period (300 days). From each run of the simulations we recorded the total number of individuals who were infected on each day in each social community. We also recorded the proportion of each social community that was concerned or adherent to social distancing on each day.

### Analysis

To compare between different runs of the simulations we quantified the height of the epidemic peak at a population level by aggregating the daily counts of symptomatic infections in all 10 communities and calculating the maximum prevalence of infected individuals (I1) in the population. This measure of the height of the epidemic peak indicated how successfully each simulated population managed to “flatten the curve” with their adherence to mitigating behaviours^[4]^. We present these results by comparing epidemic peaks when individuals learned about symptomatic network neighbours from different types of social contact while considering different values of the Social Construction and Reassurance effects.

To help explain some of the differences between the infection and communication layers in their ability to “flatten the curve” when used to provide information on local prevalence (to the Awareness effect), we also examined the similarity of connections in these layers by quantifying the proportion of contacts in each layer that were also present in the other (Fig. S1) for each of the nine multiplex networks that differed in homophily and modularity.

## Data Availability

No primary data was collected as part of this effort. All relevant code is publicly available at https://github.com/matthewsilk/CoupledDynamics2_layeruse

## Supplementary Tables and Figures

**Table S1.** Parameter values for the SEIRD model used

| Parameter | Value used |
| --- | --- |
| Transmission probability per contact | 0.3/mean degree |
| Mean length of incubation period | 5.1 days |
| Mean length of pre-symptomatic period | 6.7 days |
| Mean length of symptomatic period | 10 days |
| Mean length of hospitalisation | 4.2 days |
| Daily probability of symptomatic case becoming hospitalised for young adults and children | 0.0125 |
| Daily probability of symptomatic case becoming hospitalised for old adults | 0.025 |
| Daily probability of death in hospitalised cases for young adults | 0.012 |
| Daily probability of death in hospitalised cases for young old adults | 0.092 |

**Figure S1.**
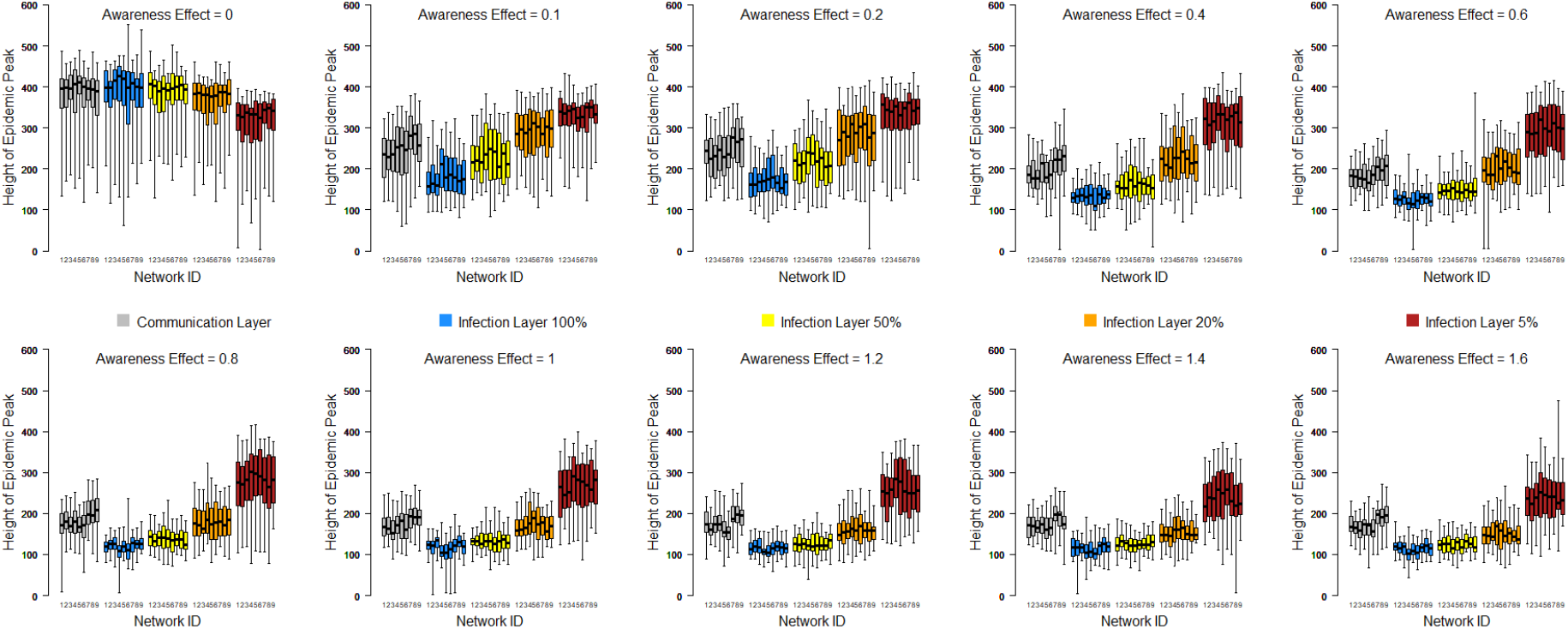
The relationship between the height of the epidemic peak and how an individual finds out about symptomatic contacts when the Social construction effect is weak (<0·3) for the full range of Awareness effects modelled and plotted separately for each of the nine multiplex networks used in the study. Networks 1-3 have no homophily in either layer, networks 4-6 have homophily in both layers and networks 7-9 have homophily in the communication layer only. In networks 1, 4 and 7 both layers have a relative modularity of 0·4, in networks 2, 5 and 8 both layers have a relative modularity of 0·6, and in networks 3, 6 and 9 the relative modularity of the infection layer is 0.6 and the relative modularity of the communication layer is 0·4.

